# Classification performance bias between training and test sets in a limited mammography dataset

**DOI:** 10.1101/2023.02.15.23285985

**Authors:** Rui Hou, Joseph Y. Lo, Jeffrey R. Marks, E. Shelley Hwang, Lars J. Grimm

## Abstract

**Objectives:** To assess the performance bias caused by sampling data into training and test sets in a mammography radiomics study.

**Methods:** Mammograms from 700 women were used to study upstaging of ductal carcinoma in situ. The dataset was repeatedly shuffled and split into training (n=400) and test cases (n=300) forty times. For each split, cross-validation was used for training, followed by an assessment of the test set. Logistic regression with regularization and support vector machine were used as the machine learning classifiers. For each split and classifier type, multiple models were created based on radiomics and/or clinical features.

**Results:** Area under the curve (AUC) performances varied considerably across the different data splits (e.g., radiomics regression model: train 0.58-0.70, test 0.59–0.73). Performances for regression models showed a tradeoff where better training led to worse testing and vice versa. Cross-validation over all cases reduced this variability, but required samples of 500+ cases to yield representative estimates of performance.

**Conclusions:** In medical imaging, clinical datasets are often limited to relatively small size. Models built from different training sets may not be representative of the whole dataset. Depending on the selected data split and model, performance bias could lead to inappropriate conclusions that might influence the clinical significance of the findings. Optimal strategies for test set selection should be developed to ensure study conclusions are appropriate.

## INTRODUCTION

Ductal carcinoma in situ (DCIS), a stage 0 form of breast cancer, accounts for about 16% of all new breast cancer diagnoses.(1) Although DCIS by itself is not life threatening (2, 3), DCIS is a potential precursor to invasive ductal carcinoma (IDC) and mixed DCIS and IDC lesions are common.(4) Furthermore, Between 10-25% of DCIS cases will be upstaged to IDC at surgery.(5, 6) Therefore, improving the pre-surgical diagnosis of DCIS and occult invasive cancer is important for treatment planning. Previous work demonstrated that clinical predictors and radiomic features extracted from mammograms can be used in machine learning models to successfully predict DCIS upstaging with receiver operating characteristic area under the curve (AUC) of 0.71.(7) Furthermore, radiomics models performed substantially better than models based on clinical features which are the current standard by which clinical decision making is based. While those studies showed strong promise, there was concern about bias in the test results which unexpectedly outperformed the training models. In the context of a difficult clinical challenge, this pattern of performance exposed the potential limitations of data sampling practices in machine learning that rely on train-test splits.

Machine learning, in particular deep learning, has shown tremendous promise to address important clinical challenges in medical imaging. However, these approaches typically require very large datasets with notable studies in chest radiographs and CT each including tens of thousands of cases for model building and testing.(8-10) In breast imaging, recent studies on mammographic lesion detection show that models can rival or even exceed the performance of expert radiologists.(11-14) Researchers must make important decisions on model design and testing which usually leads to splitting the data into subsets for training, validation, and independent testing.(15-17) With large datasets, such data splitting avoids overtraining, such that training performance will generalize to independent testing. However, many important clinical questions, such as predicting upstaging of DCIS to invasive cancer, are limited by relatively small datasets, low prevalence rates of relevant clinical outcomes, and heterogeneous confounding variables. Determining the best strategy in this setting to ensure that model results are generalizable to other new data is thus a challenging task.

In this study, we focused on the clinical task of predicting upstaging of DCIS to invasive cancer to explore the performance bias between training and test sets, and discuss options to achieve representative performance when using machine learning models to solve such classification problems. The purpose of this study was to assess the effect of different train-test sets splits on limited datasets and how they would affect actual performance and model selection.

## METHODS AND MATERIALS

### Study Population

All patients who underwent 9-gauge vacuum assisted core needle biopsy at a single health system between September 2008 and April 2017 with a diagnosis of DCIS were identified. Women aged 40 or older who presented with calcifications on digital screening mammography, without mass, asymmetry, or architectural distortion, and who had no prior history of breast cancer or DCIS were collected. The process of data collection, calcification segmentation and feature extraction have been reported in a previous study.(7) In brief, all calcifications were annotated by a fellowship-trained breast radiologist (L.J.G., with 8 years of experience), and then automatically segmented.

### Feature Extraction and Model Building

The pipeline of this study is depicted in Figure 1. 109 radiomic features and 4 clinical features were collected. Four different models were created: clinical features only, radiomics features only, clinical and radiomics features, and radiomics features with feature selection. Radiomic features described individual and clustered calcifications’ shape, texture, and topology characteristics, while the clinical features included patient age as well as DCIS estrogen receptor, progesterone receptor, and nuclear grade at diagnosis. All features were standardized with zero mean and unit variance separately for each vendor (GE or Hologic).

**Figure 1.**
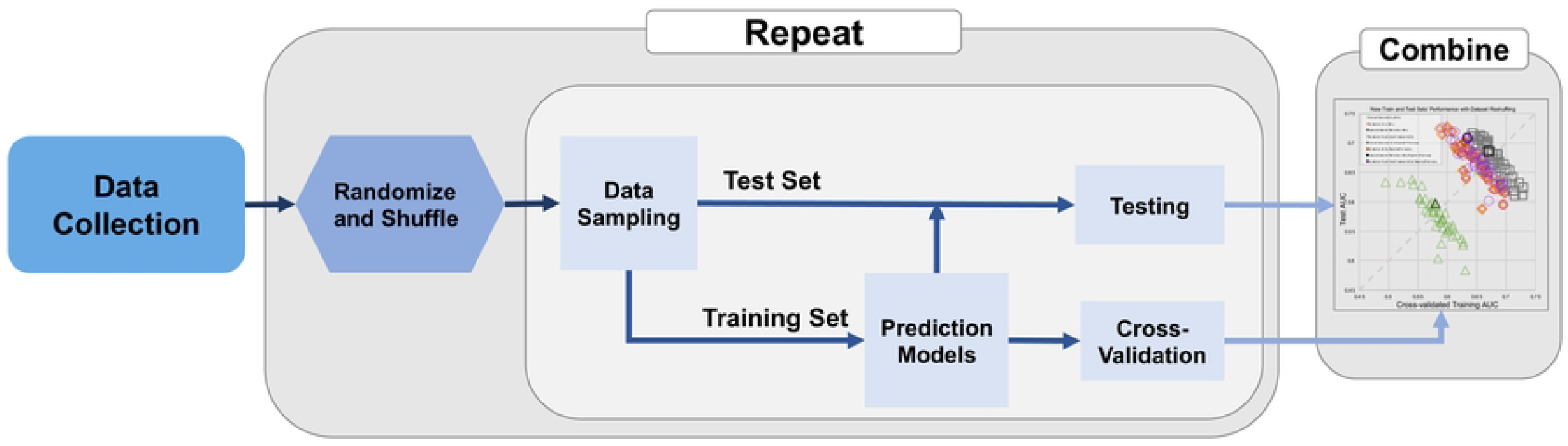
Data Resampling and Evaluation Procedure

The dataset was randomly shuffled and then divided into training and test sets while balancing the upstaging rate. Specifically, the 700 cases were composed of 586 pure DCIS and 114 upstaged DCIS (upstage rate = 16.3%). During each split, 400 DCIS cases (335 pure DCIS and 65 upstaged DCIS) were selected for training. The remaining 300 cases (251 pure DCIS and 49 upstaged DCIS) were reserved for testing.

During training, 5-fold cross-validations were repeated 200 times after randomly shuffling to alleviate effects of case ordering within the training set, and all those validation AUCs were averaged to represent the training result. For testing, the hyperparameters most frequently selected from training were applied to create a single classifier over the entire training set. To assess the effects of different data sampling, this procedure of random case shuffling, splitting into training vs. test sets, cross-validating the training set, and evaluating the test set was repeated 50 times. Each shuffle and split provided different train-test sets, thus generating a pair of training vs. test performances.

To assess whether the data sampling affected different types of classifiers, the logistic regression was replaced with support vector machines (SVM) and the procedure was repeated. Finally, all experiments above used cross-validation with a fixed number of 400 training cases. We varied the training set size and repeated cross-validations using 20 randomly sampled cohorts, while keeping the upstage rate the same. Cross-validations were performed in increments of 100 cases, adding another 100 random cases without replacement with each step until all 700 cases were used. This experiment also simulated the robustness of bypassing train-test splitting and instead just reporting cross-validation across all available cases. The entire code and features of the data will be publicly available upon publication on GitLab (https://gitlab.oit.duke.edu/railabs/LoGroup/data-resampling-bias-analysis).

## RESULTS

### Train and Test Performance with Different Shuffles

The performance of the models involving different combinations of radiomics and clinical features are shown in Figure 2. The identity diagonal defined by equal training and test performances represents perfect generalization. Each point corresponds to one train-test split, such that points below the diagonal mean that training was greater than test AUC while the opposite for holds above the diagonal.

**Figure 2.**
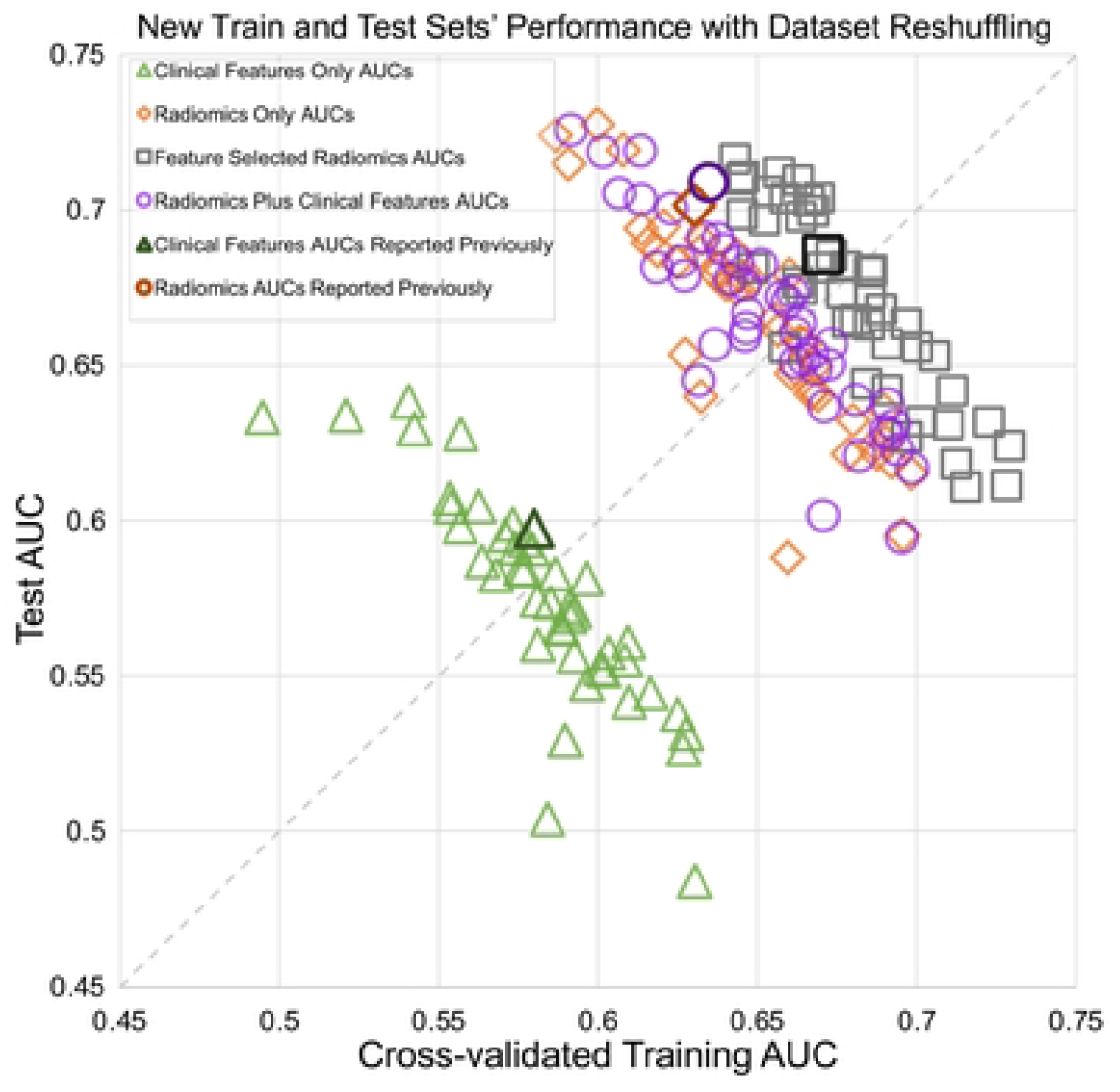
Interaction of the cross-validated training and test set performances for 4 different logistic regression model types. Scatter points were based on shuffled splits into different train-test sets. All 4 models showed anti-diagonal trend where training and test AUCs traded off against each other. Dark symbols represent a previously published study’s performances.(7)

The results for each model type (clinical and/or radiomics features) are distributed perpendicularly to the diagonal line. Training and test performances trade off against each other, i.e., higher training AUC corresponded to lower testing AUC, and vice versa. These tradeoffs created very wide ranges in both training and test AUCs: radiomic features, train 0.59-0.70, test 0.59-0.73; radiomics with feature selection, train 0.64-0.73, test 0.61-0.72; radiomics plus clinical features, train 0.59-0.70, test 0.60-0.73; and clinical features, train 0.50-0.63, test 0.48-0.64. Despite wide distributions, the first three models clustered together with similar anti-diagonal trends and performances, while the model for clinical features alone kept the same anti-diagonal trend but with lower performance. The averaged performance for each model showed different trends from results published a previous study.(7)

While shuffling and splitting, we monitored the average patient age and lesion size for training vs. test sets, as those features are established predictors of upstaging. As shown in Figure 3, during the repeated shuffles and splits for each model type, less than 10 splits yielded significant differences in age or lesion size, but those points were randomly distributed and did not show any consistent trend. In other words, even if those splits were excluded to control for these additional factors, there would be no effect on the overall distributions.

**Figure 3.**
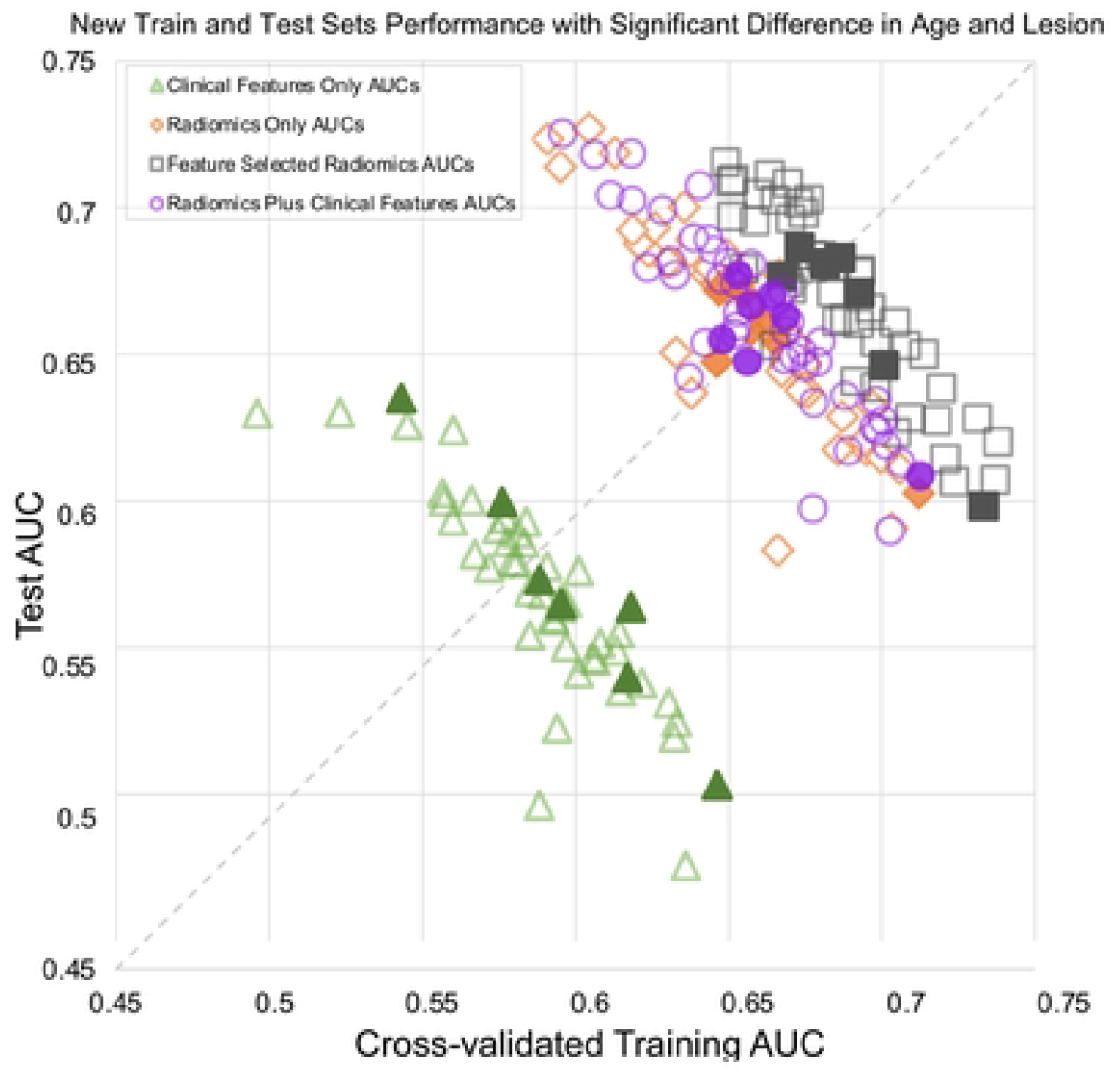
Interaction of the cross-validated training and test set performances for 4 logistic regression model types across different splits into train-test sets. Solid symbols indicate splits with significant differences in patient age or lesion size; their exclusion would not have changed the overall distributions.

### Train and Test Performance with SVM

A subset of models was selected to evaluate the effect of using a different type of classifier, the support vector machine (SVM). Hyperparameter tuning included kernel type, kernel coefficient, and regularization parameter. The model with clinical features was excluded due to the low performance, and the radiomic model with feature selection was excluded because different resulting features would preclude direct comparison. Unlike with logistic regression, the SVM performances (Figure 4) were randomly spread around the diagonal line, although there was still a wide range in AUCs: radiomic features, train 0.50-0.70, test 0.5-0.72; radiomics plus clinical features, train 0.50-0.70, test 0.48-0.71.

**Figure 4.**
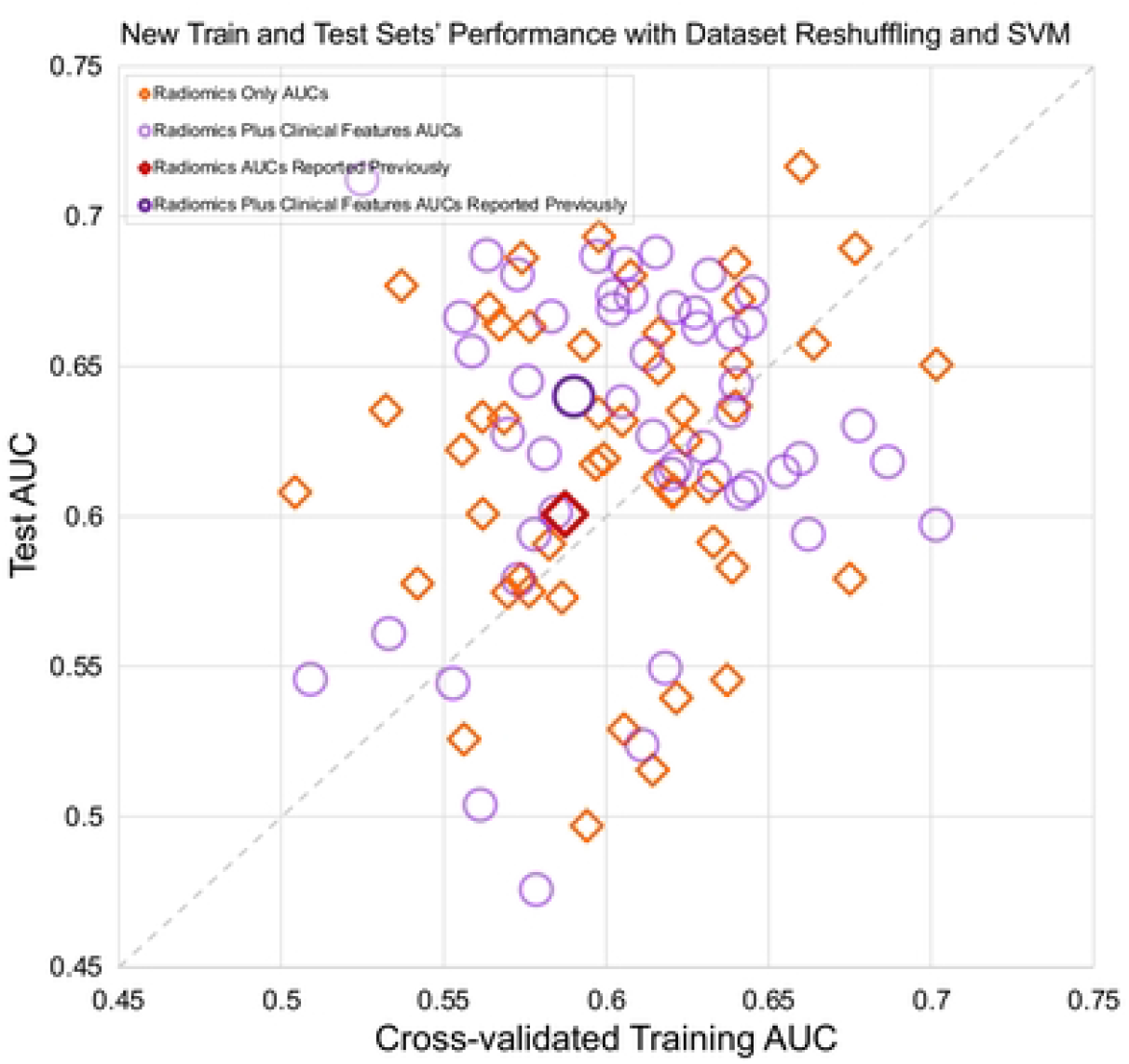
Interaction of the cross-validated training and test sets for two SVM model types. Scatter points from different train-test splits are randomly distributed.

### Cross-validation Performance with Incremental Cases

Cross-validation training performance was repeated with 20 random cohorts as the cohort size was increased using the model with radiomic features only. Boxplots of the results at each number of training cases are shown in Figure 5.

**Figure 5.**
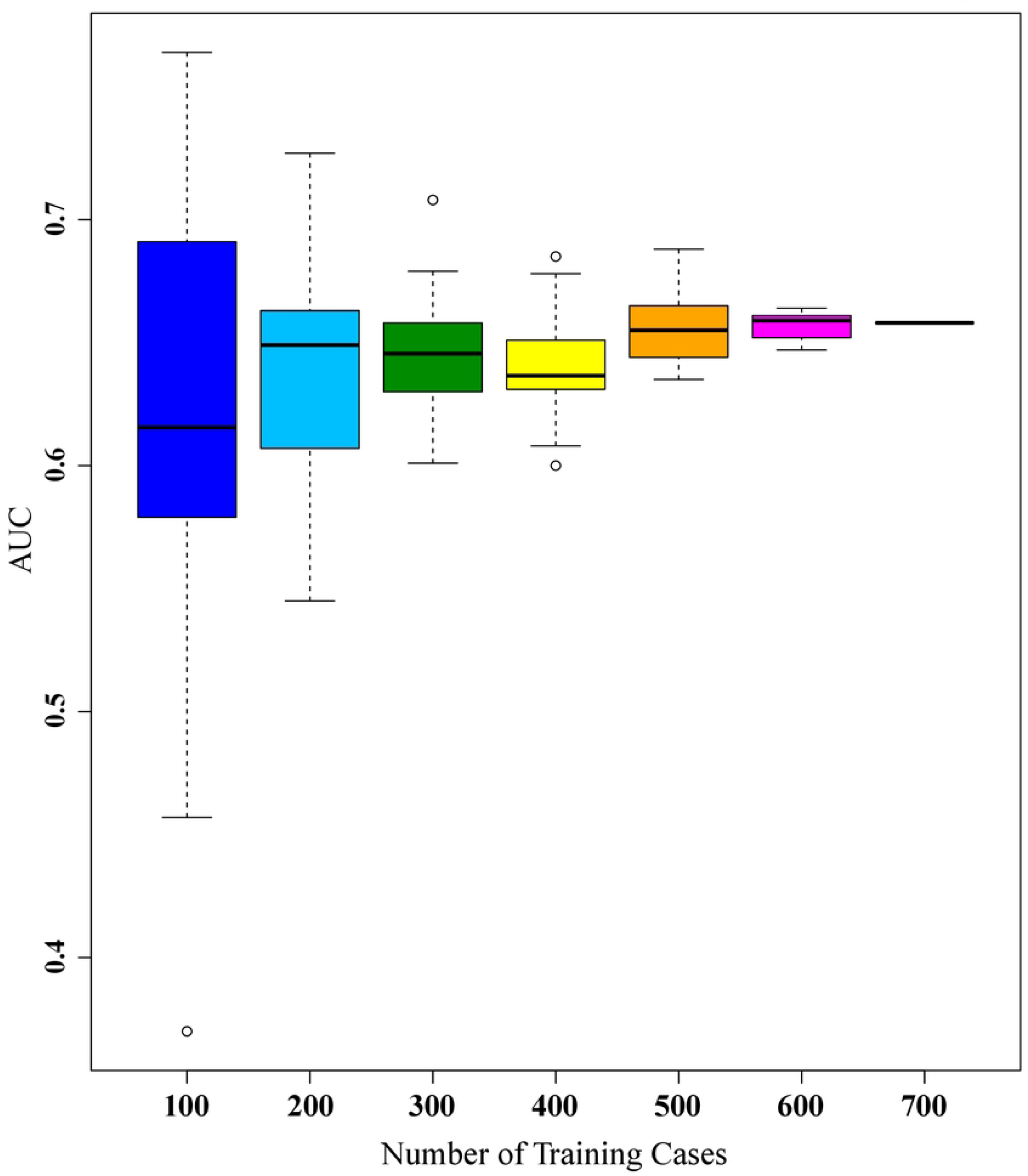
Box plots of cross-validated AUCs using different numbers of training cases.

As expected, the error bars are much larger with a smaller number of cases and narrow rapidly with increased case number. Cross-validating all 700 cases in the overall dataset generated an AUC of 0.658, which actually lies above the third quartile for our previously chosen number of 400 training cases, indicating a high likelihood of under-reporting the training performance. Likewise, the change in median values from 400 to 500 cases was comparable to the interquartile range (25^th^ to 75^th^ percentile) for 500 cases, and those median values only stopped changing after 500+ cases.

## DISCUSSION

Image data collection for medical decision-making tasks can be challenging, thus machine learning studies are often based on datasets of limited size. For example, the 700 DCIS patients in this study represents all available cases spanning almost 9 years from a large health system. Initial studies at or below this scale are important to demonstrate feasibility and justify the additional effort of larger trials or external validation. As revealed using our radiomics dataset for predicting DCIS upstaging to invasive cancer, this is an extremely difficult setting in which to generate results that are generalizable, and common methods used for sampling of limited data can cause three types of performance bias.

First, many machine learning studies utilize a one-time split of the data into training vs. test sets. This decision is motivated by the desire to reduce computational cost or to protect the test set for the final, independent assessment. Given that single split, however, the test performance is defined by one small, specific cohort of cases selected. By repeatedly shuffling and splitting, we demonstrated that the distribution of cases into training vs. test sets causes potential bias. For logistic regression, a consistent tradeoff was revealed in which higher training performance resulted in lower test performance and vice versa. Despite controlling for several key parameters (age, lesion size, and prevalence), these discrepancies were reduced only slightly.

Second, this variability caused by data sampling can affect different classifiers inconsistently. When we compared several competing models based on radiomics and/or clinical features, the different splits also randomly affected the models’ rank ordering. When the analysis was repeated for the SVM classifier, again there were often large discrepancies between training vs. test results, and their relationships were even less predictable. The aforementioned data sampling bias may have been compounded by overfitting due to the more powerful, nonlinear model.

Third, cross-validation can reduce the test variabilities by averaging performance across multiple data splits. Cross-validation over all cases is not a panacea, however, as that leaves no independent test of generalizability. We assessed this risk by repeated trials of cross-validation while varying the cohort sizes. The median performances converged to representative values only for the last few cohorts with the largest number of cases. In retrospect, cross-validated performances in previous studies based on fewer DCIS cases(18-20) represented just one sample taken from very wide distributions.

Our study has some limitations. First, the study was based on data from a single institution with a limited dataset focused on a specific clinical task. This often reflects the real-world scenarios affecting many machine learning researchers who would face similar risks for performance bias. Second, we intentionally focused most of our analyses on logistic regression because it is robust and very commonly used. For the more complex, nonlinear SVM, we only tuned common hyperparameters to show that bias between training and test sets persists for this new algorithm. With more powerful modeling techniques such as deep learning, the even greater risk for overfitting may further aggravate the bias.

In conclusion, our study demonstrates that machine learning approaches to clinical questions that involve limited datasets are at notable risk of bias. In many initial studies in radiomics or biomarkers, the number of cases and features are comparable. When limited by this “curse of dimensionality,” splitting the data further is impractical, so cross-validation over all available cases may be the only recourse, but may result in considerable bias. When there are substantially more cases than features, a single split into train-validate-test sets may suffice. Cross-validation may further reduce the bias, but confirming that requires even more data to allow the luxury of a separate test set. Paradoxically, it is difficult to confirm that there is enough data until there is enough data. To examine that uncertainty, our post hoc analyses repeated the sampling and modeling experiments hundreds of times. As a final caveat, however, such indirect but repeated exposure to all the data indirectly informs model design choices and hyperparameters, which in turn leads to optimistic bias. Ultimately, researchers should expect hidden uncertainty and bias, particularly when using relatively limited datasets. Ongoing efforts aimed at building larger and more diverse datasets are thus clearly needed to address these limitations arising from data starvation. Only with sufficiently representative data can researchers ensure reproducibility and successful clinical translation.

## Data Availability

The entire code and features of the data will be publicly available upon publication on GitLab (https://gitlab.oit.duke.edu/railabs/LoGroup/data-resampling-bias-analysis).

## Acknowledgements

Research reported in this publication was supported in part by the National Cancer Institute of the National Institutes of Health under Award Numbers U01-CA214183 and R01-CA185138, DOD Breast Cancer Research Program W81XWH-14-1-0473, Breast Cancer Research Foundation BCRF-16-183 and BCRF-17-073, Cancer Research UK and Dutch Cancer Society C38317/A24043, and an equipment grant from Nvidia Corp. The content is solely the responsibility of the authors and does not necessarily represent the official views of the National Institutes of Health.

